# Clinical and Economic Outcomes of Attention-Based Rehabilitation for Functional Neurological Disorder

**DOI:** 10.64898/2026.05.20.26353701

**Authors:** David D.G. Palmer, Stacey Palmer, Brett Darracott, Kelley Stone

## Abstract

**Introduction:** Functional neurological disorder (FND) is a common cause of neurological disability and is associated with substantial healthcare utilisation and cost. Most available treatments target specific symptom subtypes, and prospective evidence regarding the effect of treatment on health-system costs remains limited. We evaluated the real-world clinical and economic outcomes of a transdiagnostic outpatient intervention, attention-based rehabilitation (ABR).

**Methods:** We conducted a pragmatic waitlist-controlled study in 54 consecutively referred patients with neurologist-diagnosed FND attending a specialist outpatient service. Clinical outcomes—including quality of life (Short Form-36), social and occupational participation (Work and Social Adjustment Scale), symptom severity, and mental health (Hospital Anxiety and Depression Scale)—were assessed at waitlist entry, treatment commencement, treatment completion, and 6 and 12 months post-treatment. Healthcare utilisation and costs were obtained prospectively from health-service financial records for the 6 months preceding treatment, the treatment period, and two consecutive 6-month post-treatment periods. Longitudinal clinical outcomes and healthcare costs were analysed using Bayesian mixed-effects and mixture models, respectively.

**Results:** All clinical measures remained stable or worsened during the waitlist control period. Across treatment, six of eight SF-36 domains, WSAS, employment status, and both HADS subdomains improved, with maintenance through 12 months. Patient-reported symptom improvement persisted post-treatment. Expected monthly health system costs approximately halved post-treatment, with net cost savings by approximately 50 days.

**Conclusion:** A fixed-duration, symptom-agnostic outpatient ABR programme was associated with durable improvements in functioning and quality of life, alongside substantial reductions in healthcare utilisation and cost, supporting scalable symptom-agnostic treatment models for FND.

## Introduction

Functional neurological disorder is one of the most common^1^ conditions that neurologists encounter, and causes significant morbidity,^2^ unemployment, and healthcare costs.^3^ Treatments for the condition have developed substantially over the last two decades, and have become gradually more available. However, despite expanding treatment options, and clinicians’ impression of benefit, there remains a paucity of high-quality evidence for benefit, and most published interventions remain symptom-subtype specific.

The largest body of published interventions for FND target functional motor disorders. While programmes vary in duration, intensity, and composition, most employ similar physiotherapeutic techniques.^4–10^ Overall, these studies suggest benefit, although outcomes measured and follow-up durations are variable. While randomised controlled trials have been performed,^5,7,10^ the overall quality of evidence is low, and questions persist regarding generalisability, durability of benefit, and cost-effectiveness.^11^

Published approaches for treatment of functional seizures are more variable than those for functional motor symptoms. Most approaches are psychotherapies, including psychoeducation, cognitive behavioural therapy (CBT), psychodynamic interpersonal therapy (PIT), mindfulness-based therapy, and acceptance and commitment therapy (ACT),^11^ although programs more directly targeting symptoms themselves have also been described.^12,13^ As with functional motor disorders literature, evidence for benefit is suggestive, but the overall quality of evidence for these interventions is low.^11^

Evidence for treatment for functional cognitive disorder is comparatively sparse. Use of cognitive rehabilitation has been suggested,^14^ and feasibility studies of CBT^15,16^ and ACT^17^ have been published, but robust evidence for clinical effectiveness remains minimal.

Two groups have described an integrated multidisciplinary outpatient treatment for functional motor disorders, combining symptom-focused physical retraining with education and psychological support.^9,18^ These programmes suggest that coordinated, mechanism-informed treatment delivered in an outpatient setting may be feasible and effective. However, such models have largely been restricted to motor subtypes, and there remains limited evidence for transdiagnostic outpatient interventions applicable across the spectrum of FND.

With an unmet need for subtype-agnostic treatments for FND, we aimed to develop and evaluate a novel treatment model which is amenable to delivery by non-specialist teams, and can be applied to all symptoms of FND. We tested the intervention using a pragmatic waitlist-controlled trial, allowing within-patient comparison under routine clinical conditions. Given limited evidence for the effect of FND treatment on healthcare costs, we also aimed to assess the effect of our intervention on healthcare utilisation and cost.

We compared clinical and economic outcomes of our intervention—which we term attention-based rehabilitation (ABR)—across treatment and over a one-year follow-up period to a within-patient waitlist control period. We planned to include all patients treated in the first two years of operation of our clinic, however a subsequent change in funding led to modification of the treatment structure; accordingly, data collection for the present evaluation ceased at 21 months. We present the clinical and economic outcomes from this evaluation period here.

## Methods

### Design

We conducted a waitlist-controlled trial, allowing within-participant comparison of outcome trajectories. Comparisons were made between changes in outcomes across the period that participants were on the waiting list for treatment and the period from the start of treatment to 12 months after finishing treatment (including both six-and twelve-month post-treatment timepoints).

### Participants

All patients referred to our treatment clinic were eligible for enrolment. The programme accepted referrals for people aged 16 and older with FND diagnosed by a neurologist and who had been informed of the diagnosis. Comorbidities and lack of agreement with the diagnosis were not considered contraindications to treatment so long as patients were willing to attend and engage in the treatment. Consent to participate in data collection did not affect patients’ acceptance into the treatment programme. The data of patients for whom the diagnosis of FND was withdrawn after initial assessment, or who declined to participate in treatment were excluded from analysis. The addition of comorbid diagnoses, or poor adherence by patients who accepted treatment were not, however, exclusion criteria. Patients were treated in the order they were referred.

### Intervention

We developed an integrated outpatient treatment model designed to treat FND regardless of symptom subtype. The intervention comprised six, one-hour treatment sessions, spaced two weeks apart, for a total duration of 10 weeks. Central to the programme was a set of techniques we refer to as attention-based rehabilitation (ABR). ABR extrapolates techniques already used in FND rehabilitation, focussing on helping patients recognise the role of symptom-focused attention in maintaining or precipitating symptoms, either during symptom expression (as in functional motor symptoms) or in the prodromal phase (as in functional seizures), and employing the converse of this to control the symptoms. Patients are taught to identify these processes and to apply strategies that require the conscious direction of attention to a competing task. With practice, patients become more proficient at this, leading in the short term to voluntary control of symptoms, and in the long-term to deconditioning of the symptoms with eventual remission. For episodic symptoms where patients are not already aware of prodromal symptoms (the majority), an additional first step of training patients to identify their prodromal symptoms is added. The programme included only limited treatment of psychological comorbidities, and no direct assessment for, or treatment of trauma. Patients were given the expectation that by the end of treatment they should have the skills required to continue improving, even if they had not achieved remission. The form of ABR is more comprehensively discussed in an upcoming article.

### Data Collection

Because FND is heterogeneous, no single outcome measure captures meaningful clinical change.^19^ We therefore measured quality of life, social and occupational participation, and symptom severity as co-primary outcomes. The primary economic outcome measure was the change in expected per-patient health service expenditure. Secondary outcomes were severity of symptoms of depression, severity of symptoms of anxiety, and expected number of encounters with individual components of the health service. Outcome measures were chosen with reference to published guidance on outcome measurement in FND.^19^

Participants were asked to complete a battery of online questionnaires at the time they were placed on the waitlist, in the week before starting treatment, at the completion of treatment, and six- and twelve-months after the end of treatment. Questionnaires were initially administered using Microsoft Forms, with a change of platform to REDCap^20,21^ approximately mid-way through the study. Questionnaires included demographics, year of onset and year of diagnosis of FND, symptoms of FND that the person had ever experienced, symptoms of FND that the person had experienced in the four weeks preceding the questionnaire, and work status. Clinical measures included in the questionnaires were the Short Form-36 (SF-36),^22^ an eight-domain measure of quality of life; Work and Social Adjustment Scale (WSAS),^23^ a five item measure of ability to engage in social and occupational activities; Patient Global Impression of Severity (PGI-S) and Patient Global Impression of Change (PGI-C),^24^ Likert-style questions assessing symptom-severity and change in symptoms; and the Hospital Anxiety and Depression Scale (HADS),^25^ which measures symptoms of anxiety and depression.

### Clinical Data Analysis

Data were analysed using Python, with Bayesian regressions performed using the Bambi package.^26^ Demographics and baseline data are summarised as mean (SD) or median (IQR), as appropriate.

Longitudinal outcomes were analysed using Bayesian piecewise generalised linear mixed-effects models, with fixed effects with participant-level random slopes for time since enrolment and time since treatment completion, a level change at the end-of-treatment time point, and participant-level random intercepts. Continuous bounded outcomes (e.g. clinical scores) were modelled using a beta likelihood with a logit link function, and ordinal categorical outcomes were modelled using cumulative logit models. Weakly informative priors were used for all models. To prevent undue influence of boundary values (0 and maximum scores) on beta regression estimates, bounded outcomes were transformed prior to modelling using the adjustment proposed by Smithson and Verkuilen ^27^ (equation 1).

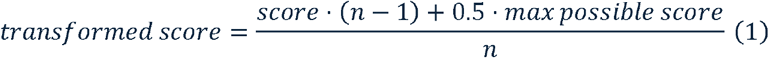

Results are presented as posterior means with 95% highest density intervals (HDIs) for model-implied estimates at each timepoint. To assess the immediate and ongoing effects of treatment relative to the waitlist control period, posterior distributions of the baseline time slope, the treatment-associated level change, and overall time slope for the follow-up period (the sum of the baseline and post-treatment slopes) were also examined with respect to zero. These effects are interpreted using regions of practical equivalence (ROPEs), representing outcomes of no clinically meaningful change. Where available, ROPE boundaries were set using published minimum clinically important differences (MCIDs). Where MCIDs were unavailable, boundaries were defined as an effect size of +/-0.1 standardised mean differences on the model scale.

Strength of evidence was summarised using posterior odds, calculated by comparing the posterior probability mass within each of three regions (within the ROPE, above the ROPE upper bound, and below the ROPE lower bound) to its complement, with interpretation guided by the updated Jeffreys scale described by Lee and Wagenmakers.^28^

Anonymised data and analysis scripts are available are available from the corresponding author upon reasonable request.

### Economic Data Analysis

Economic analysis was performed with reference to CHEERS guidelines. Costs were analysed from the health system perspective in contemporary Australian Dollars with no discounting, given the short timeframe over which data were gathered. Data on healthcare encounters and costs for all facilities within SCH were collected directly from the health service patient costing system, ensuring complete data were available for all participants. Data were collected for the six-month period preceding the start of treatment, the treatment period, and two consecutive six-month periods following the end of treatment. The six-month pre-treatment window was used regardless of the timing of onset of each patient’s FND, or the timing of the referral for treatment.

Healthcare costs were analysed at the patient–period level using a Bayesian mixture model designed to estimate expected health-system expenditure for the treated cohort. Cost data were extremely right-skewed, with a small number of patients accounting for a large proportion of total spend, and a substantial proportion of patients having zero cost in post-treatment periods. To accommodate this structure, costs were modelled using a three-component mixture likelihood comprising (i) a point mass at zero to represent periods with no healthcare utilisation, (ii) a “typical cost” component, and (iii) a “catastrophic cost” component, representing rare periods of exceptionally high expenditure, both modelled using log-normal distributions. The mixture weights were parameterised such that the probability of any cost (Ψ) being incurred and the probability that a cost arose from the typical versus catastrophic component (ω) were allowed to vary by study period. Positive costs were modelled on a per-day log scale with a log-exposure offset for the length of each observation window, allowing estimation of total costs over unequal follow-up periods. To ensure identifiability, the mean of the catastrophic component was constrained to exceed that of the typical component. The primary estimand was the expected total healthcare spend for the cohort in each period, obtained by integrating over the full posterior distribution, including zero-cost and high-cost observations. Cost savings were quantified as posterior differences in expected cohort-level expenditure relative to the pre-treatment period, with uncertainty summarised using 95% highest density intervals and posterior probabilities of cost reduction.

Encounters with individual sections of the health service (emergency department, inpatient care, and outpatient care) in each period were compared using negative binomial regression, with the primary estimand the expected number of presentations per patient per period. ROPEs for these comparisons were set as one tenth of the first of each pair of compared values.

## Results

67 patients attended the clinic during the study period. Of these, 62 gave consent to participate in the study (Figure 1). The data for five patients who attended one appointment and elected not to proceed with treatment, and one patient for whom the diagnosis of FND was revised, were excluded from analysis. Of the 54 patients included in analysis, 41 answered at least one questionnaire. Clinical review (DP, KS) found no clear relationship between questionnaire response and clinical features, or subjective clinician-impression of treatment outcome.

**Figure 1:**
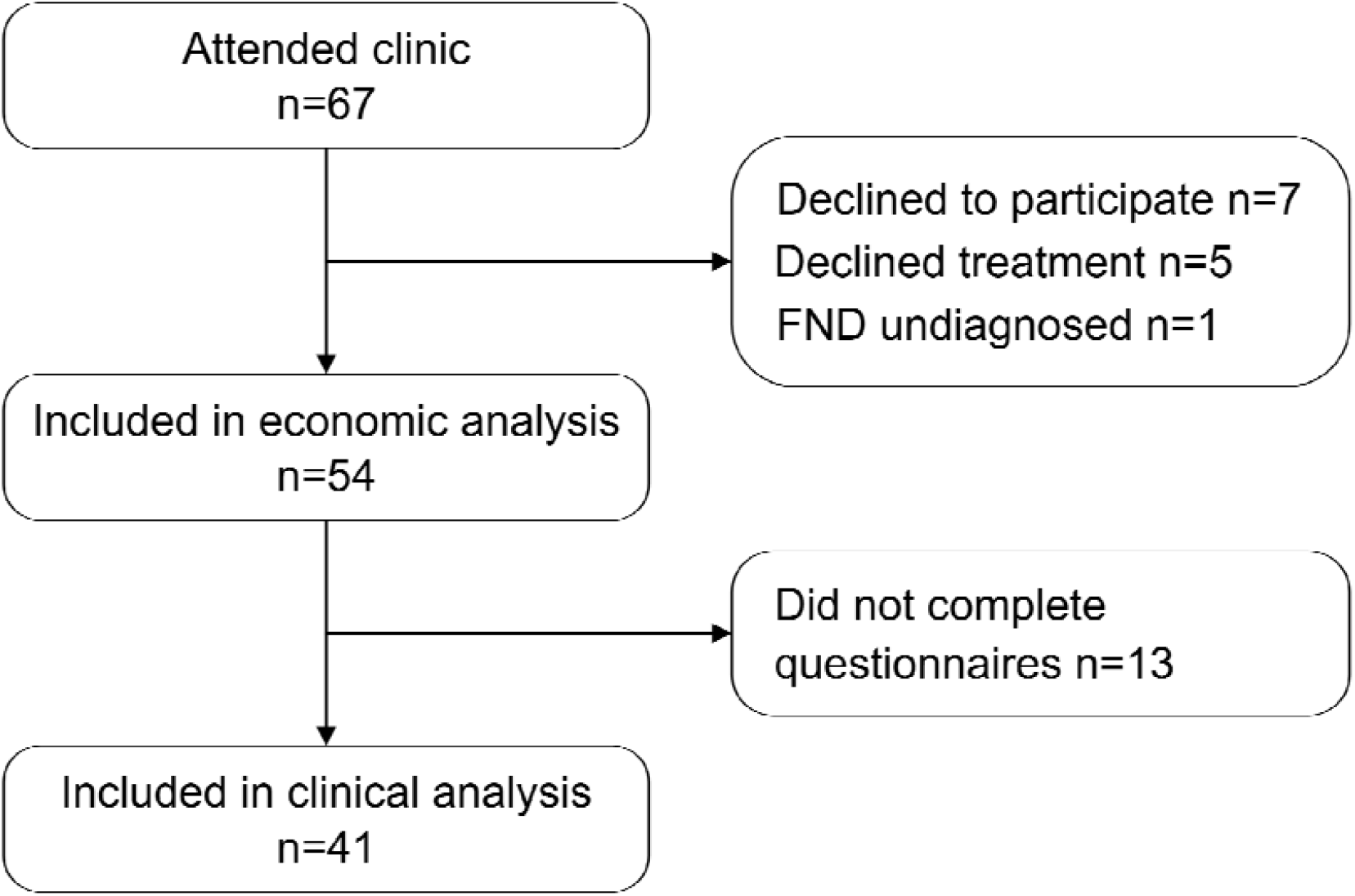
Participant flow

The mean time from enrolment on the waitlist to the start of treatment was 148 days. Demographics of participants who answered at least one questionnaire are presented in Table 1. The median FND symptom duration at enrolment for participants who responded to at least one questionnaire was 804 days (IQR 150 – 3296 days). For clinical follow-up, the mean time in the waitlist control condition was 148 days (SD 71 days).

**Table 1:**
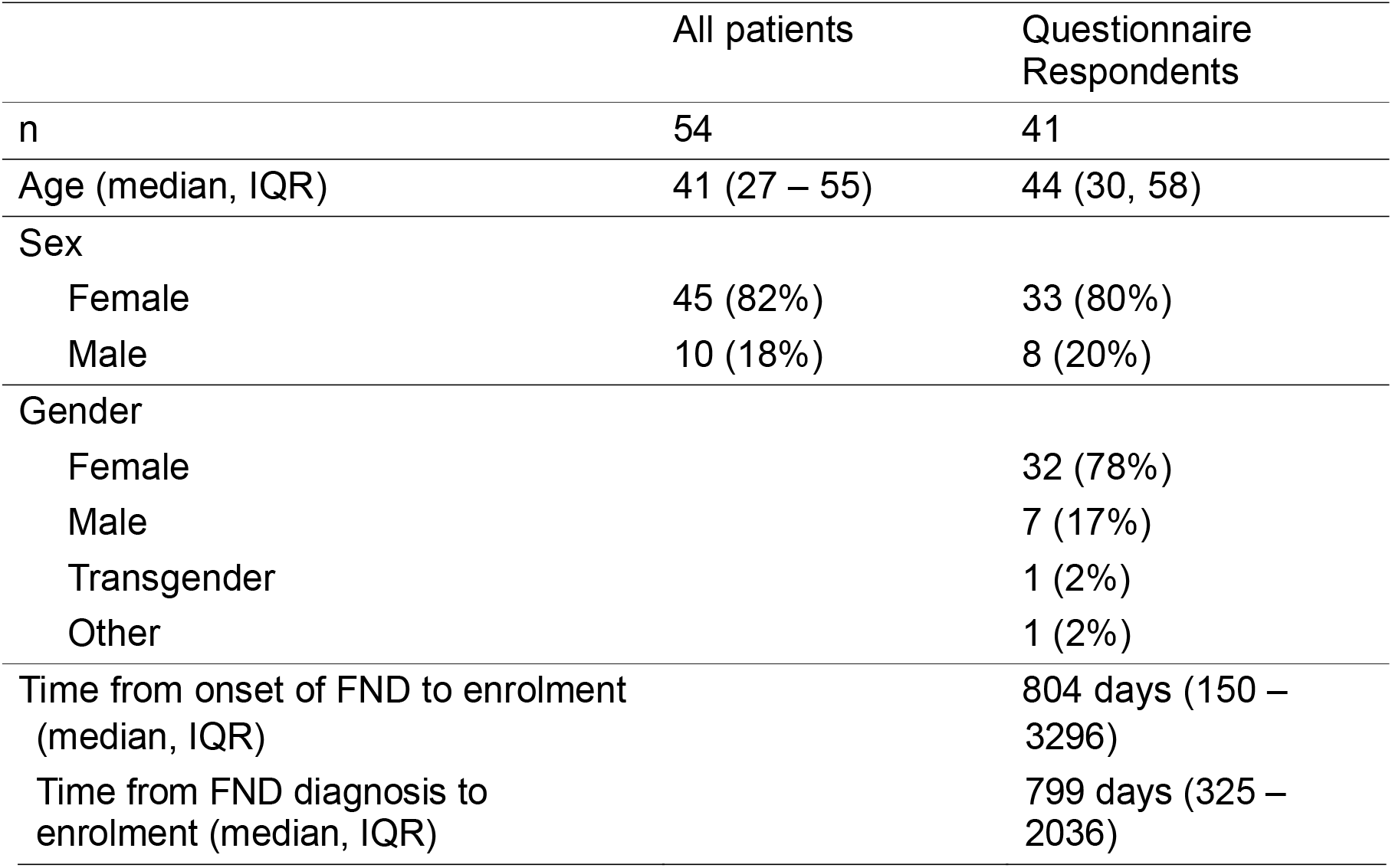
Participant demographics.

### Clinical Outcomes

Quality of life was assessed using the SF-36. Results for the eight subdomains are presented in Figure 2. For the waitlist control period, strong evidence or greater was seen for no change with respect to the MCID^29^ for physical role limitation, energy, emotional wellbeing, and general health. For all subscales, the evidence was strongest for no change. Across the treatment period, strong or greater evidence was seen for improvement in scores for physical role limitation, physical functioning, emotional role limitation, energy, emotional wellbeing, and pain. In the follow-up period, the evidence was strongest for no change on all subscales, but this was only strong or greater for the general health subscale. Note should be made that evidence for ‘no change’ reflects posterior mass within the pre-specified ROPE, rather than acceptance of a null hypothesis.

**Figure 2:**
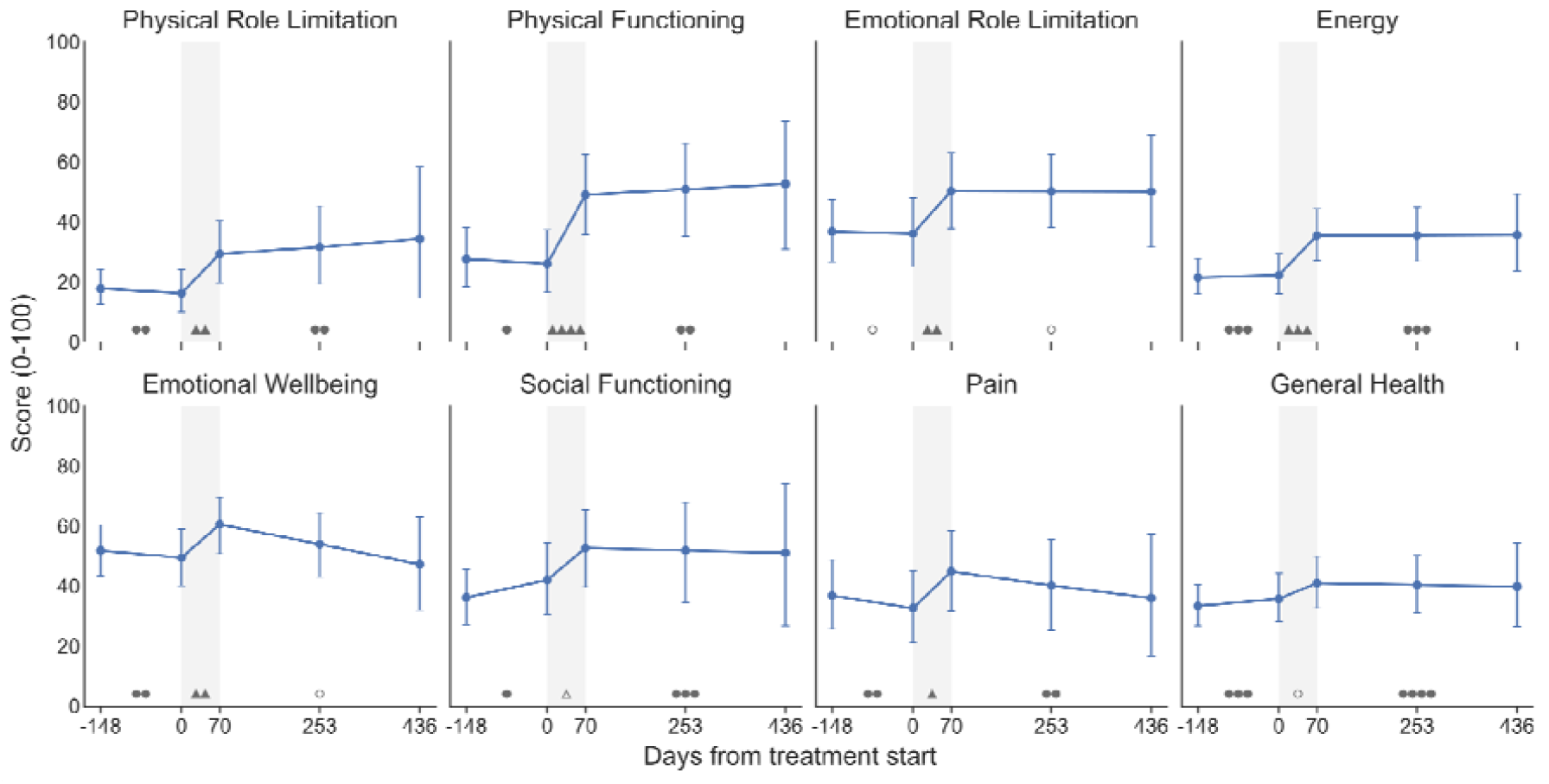
Short Form-36 subdomain scores over the study period. Points show posterior means with error bars denoting 95% highest density intervals (HDIs). The treatment period is shaded in grey. Symbols summarise Bayesian evidence for change during the waitlist, treatment, and follow-up periods. Direction of change is encoded by symbol shape (circles indicate no meaningful change), and strength of evidence by symbol count (unfilled = weak; one = moderate; two = strong; three = very strong; four = extreme evidence).

Regression results for the WSAS are presented in Figure 3. Participants were severely impaired at baseline, with the mean WSAS score 29 of a possible 40. There was extreme evidence for no change with respect to the MCID^30^ in the waiting list control period, very strong evidence for improvement (mean 8 points) across the treatment period, and strong evidence for no change across the follow-up period.

**Figure 3:**
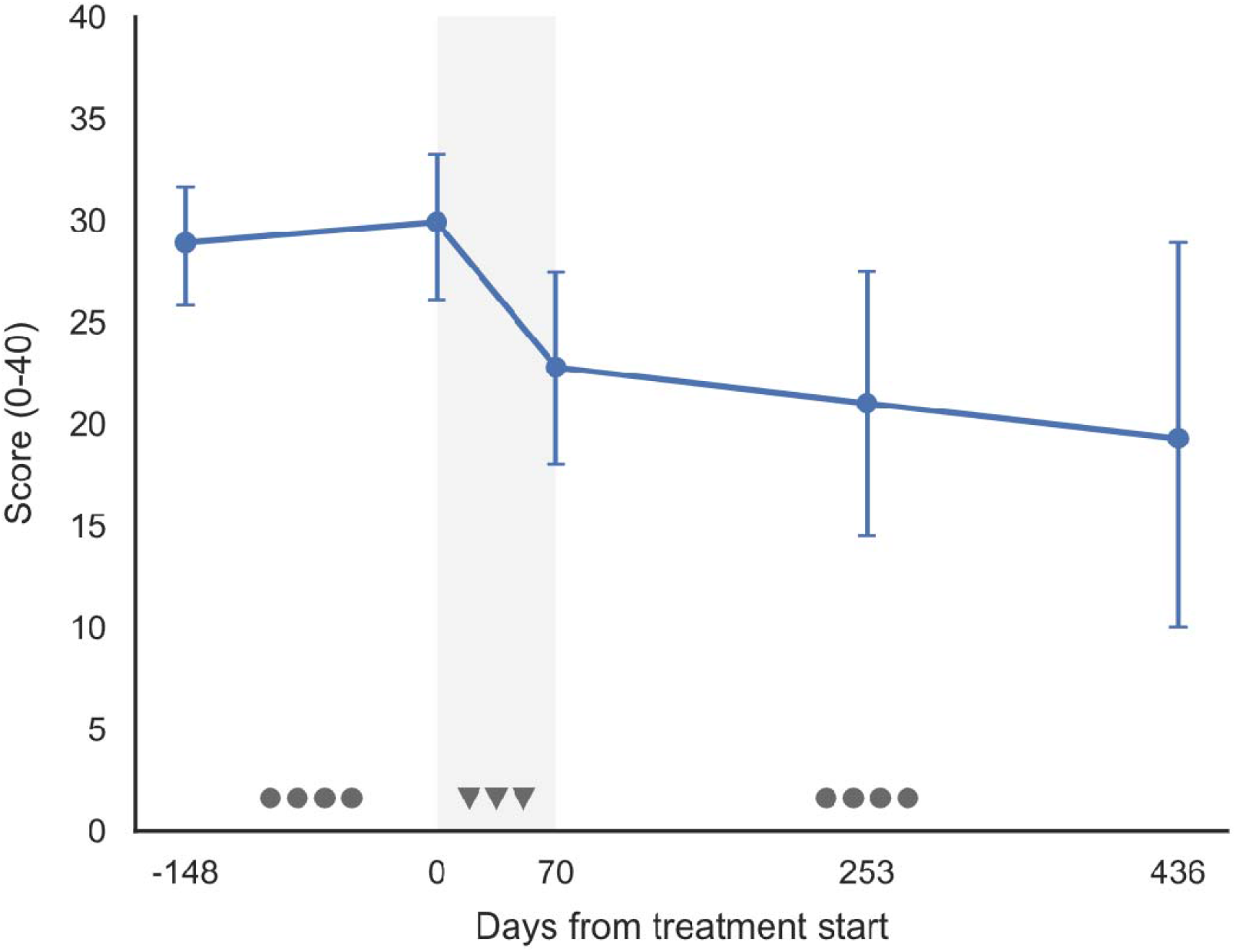
Work and Social Adjustment Scale scores over the study period. Points show posterior means with error bars denoting 95% highest density intervals (HDIs). The treatment period is shaded in grey. Symbols summarise Bayesian evidence for change during the waitlist, treatment, and follow-up periods. Direction of change is encoded by symbol shape (circles indicate no meaningful change), and strength of evidence by symbol count (unfilled = weak; one = moderate; two = strong; three = very strong; four = extreme evidence).

Through PGI-C responses, patients were somewhat more likely to report improvement than worsening of symptoms at the end of the waitlist control period (Figure 4). At the end of treatment and both follow-up timepoints, a large majority of patients rated their symptoms as improved, with the modal answer ‘much improved’. There was strong evidence (posterior odds=19) for the improvements in rating seen across the treatment period, and no meaningful evidence for change or for no change over the follow-up period.

**Figure 4:**
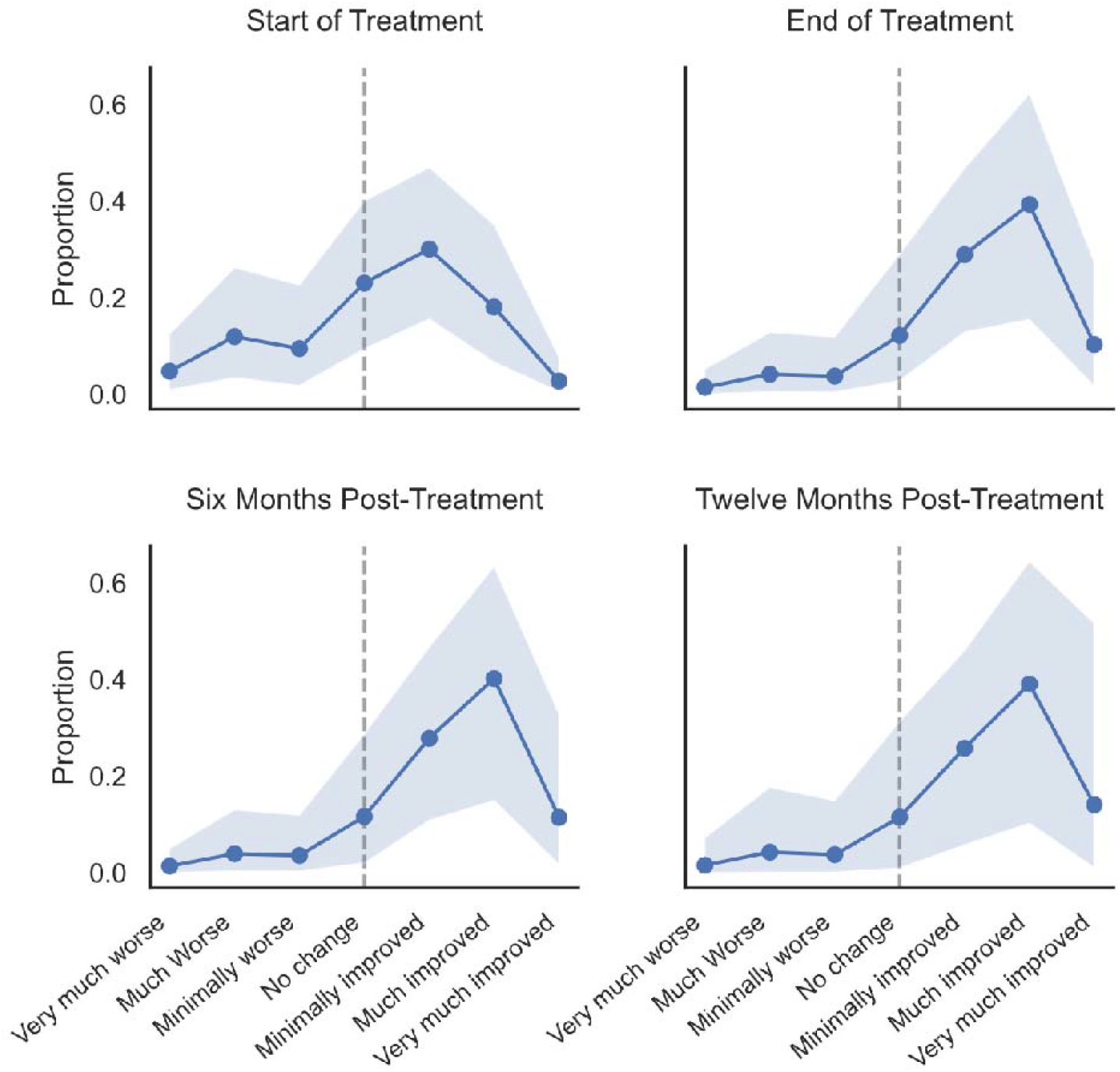
Expected distributions for Patient Global Impression of change (PGI-C) over the post-treatment follow-up period. Points show posterior means for the proportion giving each response, with shaded areas representing 95% HDI.

For both the anxiety and depression subscales of the HADS (Figure 5), weak and moderate evidence respectively was seen for a worsening in score beyond the MCID^31^ in the waitlist control period, with extreme evidence seen for improvement in score across the treatment period, and weak and moderate evidence respectively seen for continued improvement in the follow-up period.

**Figure 5:**
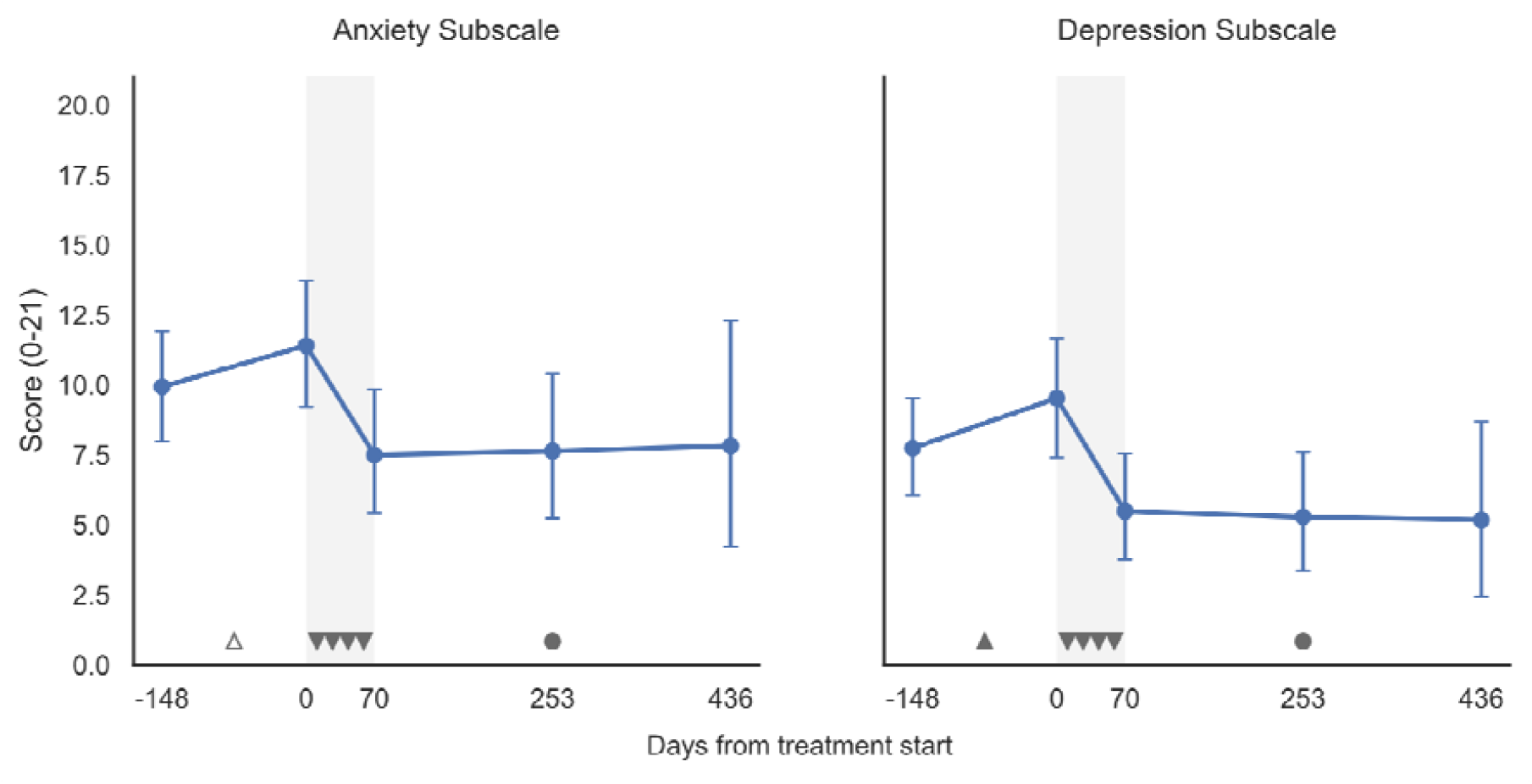
Hospital Anxiety and Depression Scale scores over the study period. Points show posterior means with error bars denoting 95% highest density intervals (HDIs). The treatment period is shaded in grey. Symbols summarise Bayesian evidence for change during the waitlist, treatment, and follow-up periods. Direction of change is encoded by symbol shape (circles indicate no meaningful change), and strength of evidence by symbol count (unfilled = weak; one = moderate; two = strong; three = very strong; four = extreme evidence).

Distributions of engagement in paid employment for patients who were not unemployed by choice (eg retired) are presented in Figure 6. For these patients, there was very strong evidence (posterior odds = 32) for worsening in work status while in the waitlist control condition, with extreme evidence for improvement in work status across the treatment phase, and moderate evidence for improvement over the follow up period.

**Figure 6:**
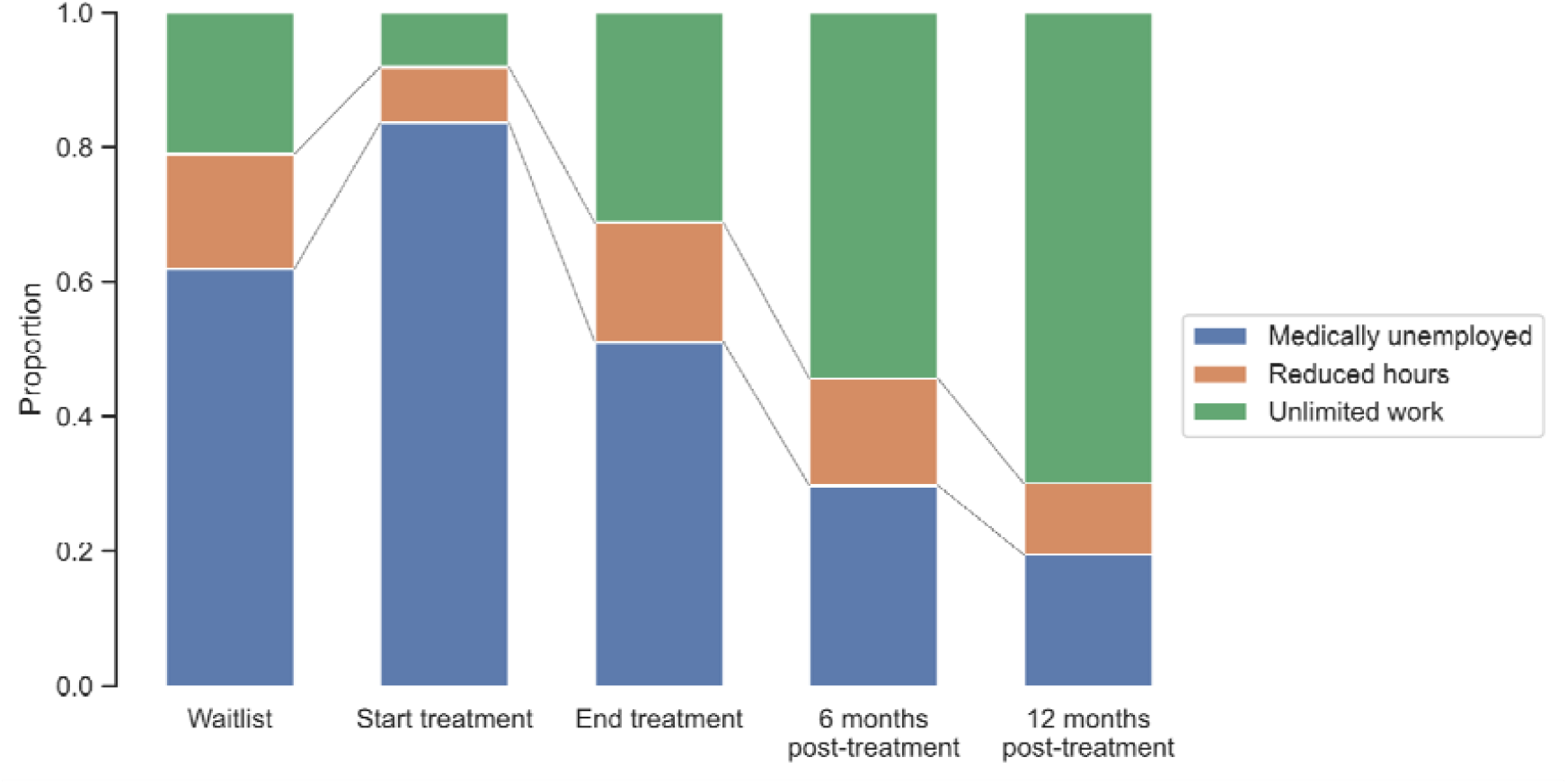
Work status over the study period. Bar size represents posterior mean for proportions of participants with each work status. Symbols summarise Bayesian evidence for change during the waitlist, treatment, and follow-up periods. Direction of change is encoded by symbol shape (circles indicate no meaningful change), and strength of evidence by symbol count (unfilled = weak; one = moderate; two = strong; three = very strong; four = extreme evidence).

### Economic Outcomes

Since health service use data were collected directly from the health service patient costing system, complete data were available for all participants. Figure 7 panel A presents the expected cost per patient per month based on the lognormal mixture model. After increasing during the treatment period, expected monthly health service expenditure per patient approximately halved for both the six and 6—12-month follow-up periods, with strong, and extreme evidence respectively for reduction in cost compared to the pre-treatment period. Figure 7 panel B presents the expected cumulative healthcare expenditure per patient compared to the counterfactual, derived by extrapolating forward the pre-treatment rate of expenditure. Based on this projection, the mean time to net cost saving after treatment is 49 days post-treatment.

**Figure 7:**
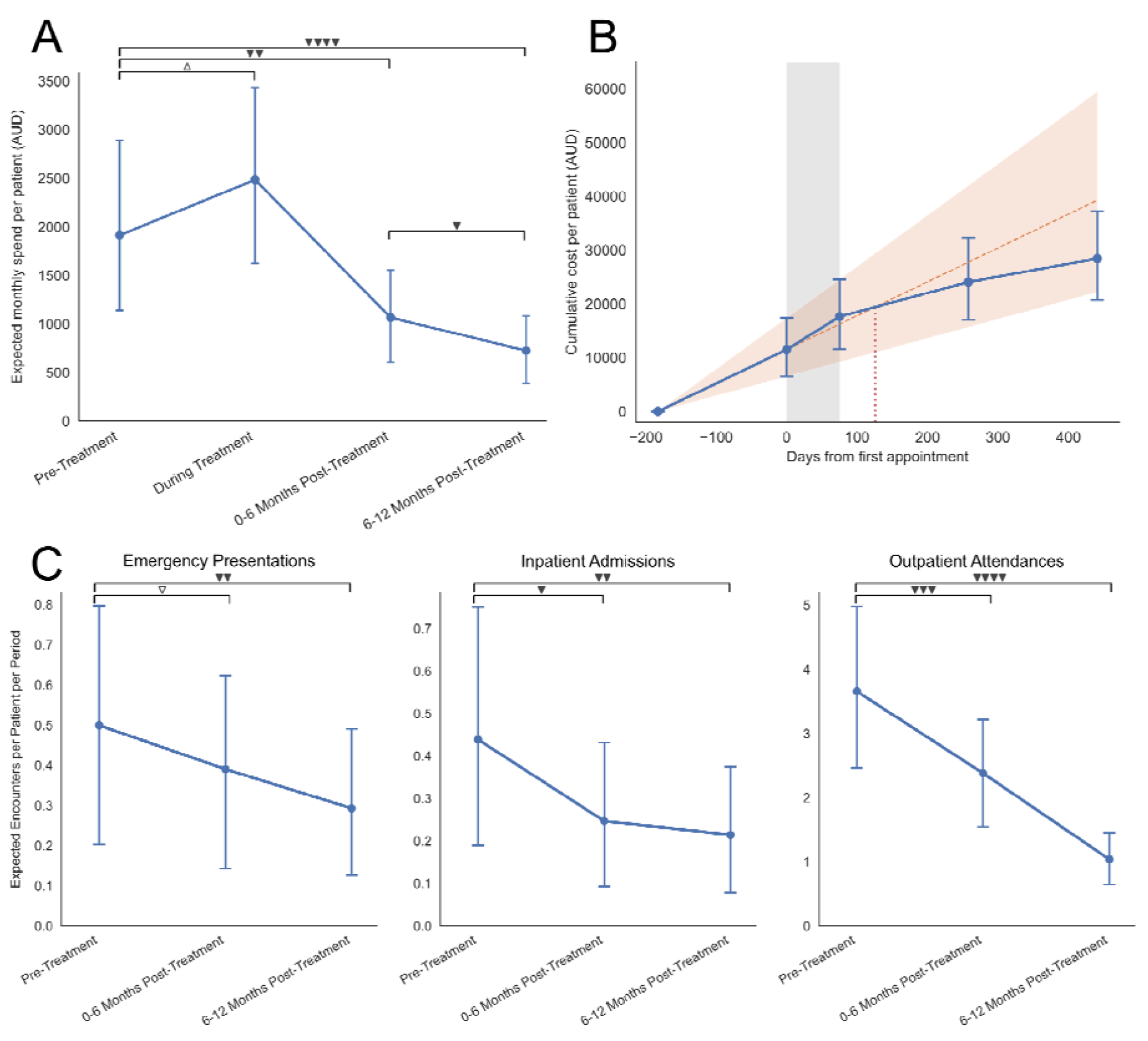
**A: Expected healthcare expenditure per patient per month in each period.** Points show posterior means with error bars denoting 95% highest density intervals (HDIs). Symbols summarise Bayesian evidence for change during the waitlist, treatment, and follow-up periods. Direction of change is encoded by symbol shape (circles indicate no meaningful change), and strength of evidence by symbol count (unfilled = weak; one = moderate; two = strong; three = very strong; four = extreme evidence). **B: Cumulative healthcare cost per patient compared to the counterfactual of no treatment**. Blue points and error bars indicate mean and 95% HDIs for actual cumulative expenditure. Dashed orange line and shaded area represent the linear extrapolation of expenditure from the waitlist control period. The dotted purple line indicates the posterior mean time to cost neutrality (day 49 post-treatment). **C: Expected number of healthcare encounters per patient per six-month period**. Mean and 95% HDI with direction with evidence for change encoded by symbols as in panel A.

Figure 7 panel C presents the expected number of healthcare encounters for each subdivision of the health service. After treatment, the expected number of emergency department presentations, inpatient admissions, and outpatient attendances per patient reduced, with weak, moderate, and very strong evidence respectively for a decrease greater than the ROPE. Between the six-month post treatment and 6-12 months post-treatment timepoints, moderate, weak and extreme evidence were seen for continued decrease in attendances. Comparison of the pre-treatment and 6-12 month follow-up periods showed strong or greater evidence for decrease in encounters in all categories.

## Discussion

Our results show that treatment with ABR was associated with improvement in each of our co-primary clinical endpoints in people with FND: quality of life, social and occupational participation, and severity of symptoms. Treatment was associated with improvement in six of eight domains of quality of life measured by the SF-36, and in all but one of those categories, there was strong or greater evidence for maintenance of the improvements across the follow-up period. Similarly, WSAS scores improved across the treatment period, with very strong evidence for maintenance of this improvement across the follow-up period. Finally, patients reported improved symptoms after treatment, which was maintained throughout follow-up period.

In addition to the improvements in clinical status, our results suggest that ABR is associated with improvement in our primary economic endpoint, expected health service expenditure. Compared to the waitlist control period, expected monthly expenditure per patient approximately halved after treatment, with further reduction in the 6—12 month follow-up period. Comparison to the counterfactual obtained by extrapolating forward costs from the waitlist control period suggests that the provision of the intervention results in net cost saving to the health service approximately 50 days after the completion of treatment.

An important feature of this study is the duration of follow-up. Few published interventions for FND report outcomes beyond six months after completion of treatment, and sustained follow-up to 12 months or longer remains uncommon. Although direct comparison across studies is limited by heterogeneity in patient populations and outcome measures, the magnitude and durability of clinical improvement observed in this study appear comparable to, and in some domains greater than, those reported in the most effective programmes described to date. Notably, these outcomes were achieved with a fixed-duration outpatient intervention of moderate intensity, supporting the potential for durable benefit without prolonged or resource-intensive treatment.

Important results were also seen for our secondary endpoints. Engagement in paid employment increased across the treatment period after worsening during the waitlist control period. Similarly, symptoms of anxiety and depression, which worsened during the waitlist control period, improved dramatically across treatment, with maintenance of improvement through the follow-up period. The latter finding is particularly notable for the fact that our intervention is explicit in not directly treating mental health disorders (although psychological techniques which may be helpful are taught), suggesting that improvement in FND symptoms may, for some patients, contribute to secondary improvement in mental health.

The inclusion of waitlist control and follow-up periods in our study allows substantial confidence in our results. No measures improved across the waitlist control period, and some worsened, making it unlikely that regression to the mean accounts for the improvements associated with treatment. Furthermore, the addition of follow-up timepoints extending to 12 months after the end of treatment increases our confidence that the improvements in outcomes associated with ABR are enduring, consistent with our goal of building skills for self-treatment.

While a wealth of evidence exists for increased healthcare costs associated with FND,^3^ few studies have assessed the effect of a treatment on costs. Our economic analysis is consistent with marked cost savings after treatment with ABR, driven by reduced emergency presentations, inpatient admissions, and outpatient visits. While it is moderately resource intensive in the short term, these results show that provision of the intervention by health services can be expected to lead to net reductions in cost less than two months after treatment is completed, with cost savings enduring to at least one year after completion of treatment.

Beyond its effectiveness, we believe that this model of care has significant strengths, which could enable wider access to effective FND treatment. As a subtype-agnostic intervention, this treatment can be applied to all symptoms of FND, obviating the need for duplication of FND treatment services for different symptoms. The model of care, with three clinicians working together in the same room also supports rapid trans-disciplinary learning, potentially enabling clinicians without prior subspecialist experience to deliver the intervention, meaning clinics could be developed broadly without the need for delivery by clinicians subspecialised in treating FND. Its fixed-length design allows predictable throughput and limits indefinite continuation of resource-intensive treatment. Finally, we hope our demonstration of its strong economic benefits aids in arguments for increased funding for FND treatment, expanding availability.

Our study has limitations. Given its waitlist-controlled design, blinding of outcomes was not possible. This design is also less effective at mitigating the effect of confounders than a randomised controlled trial. As is frequently the case with longitudinal studies using patient-reported outcomes, our clinical data has substantial missingness. While clinical review did not reveal an obvious pattern to the missingness, the possibility that the missing data are missing not at random (MNAR), and might consequently have introduced systematic bias, cannot be excluded. However, economic outcomes were complete for all participants, partially mitigating concerns about attrition bias. Finally, while we believe this treatment is likely to be appropriate for clinicians inexperienced in FND treatment to adopt, the single centre design of our study with a single clinical team limits generalisability in interpretation of the results.

In summary, our results show strong evidence of both meaningful improvement in multiple clinical domains, and health service level cost savings associated with provision of a structured ABR programme for patients with FND, regardless of FND symptom subtype. Further studies assessing the effect of implementation by other teams in other settings will further strengthen confidence in the approach.

## Author Contributions

David D. G. Palmer: Conceptualization, Data curation, Formal analysis, Funding acquisition, Investigation, Methodology, Visualization, Writing - Original Draft

Stacey Palmer: Conceptualization, Investigation, Writing - Review & Editing

Brett Darracott: Data curation, Formal analysis, Writing - Review & Editing

Kelley Stone: Conceptualization, Data curation, Investigation, Methodology, Project administration, Writing - Review & Editing;

## Ethics Statement

This work received ethical exemption by the Metro North Human Research Ethics Committee B with reference HREC/2023/MNHB/97450, in accordance with the National Statement on Ethical Conduct in Human Research (Australia). Informed consent was obtained from all participants.

